# Efficacy and Acceptability Comparisons of Cognitive Behavior Therapy, Drugs, and Their Combination for Panic Disorder in Adults: a Network Meta-analysis

**DOI:** 10.1101/2020.02.27.20028605

**Authors:** Fengjie Gao, Hairong He, Bin Yan, Jian Yang, Yajuan Fan, Binbin Zhao, Xiaoyan He, Qingyan Ma, Baijia Li, Yuan Gao, Li Qian, Zai Yang, Ce Chen, Yunchun Chen, Chengge Gao, Feng Zhu, Wei Wang, Xiancang Ma

**Affiliations:** Department of Psychiatry, The First Affiliated Hospital of Xi’an Jiaotong University, 277 Yanta West Road, 710061 Xi’an, Shaanxi, China; Clinical Research Center, The First Affiliated Hospital of Xi’an Jiaotong University, 277 Yanta West Road, 710061 Xi’an, Shaanxi, China; Center for Brain Science, The First Affiliated Hospital of Xi’an Jiaotong University, 277 Yanta West Road, 710061 Xi’an, Shaanxi, China; Clinical Research center for Psychiatric Medicine of Shaanxi Province, The First Affiliated Hospital of Xi’an Jiaotong University, 277 Yanta West Road, 710061 Xi’an, Shaanxi, China; Center for Translational Medicine, The First Affiliated Hospital of Xi’an Jiaotong University, 277 Yanta West Road, 710061 Xi’an, Shaanxi, China

**Author notes:** Address correspondence to Dr. Wei Wang at Department of Psychiatry and Dr. Xiancang Ma at Department of Psychiatry; Clinical Research center for Mental Disease of Shaanxi Province; Center for Brain Science, The First Affiliated Hospital of Xi’an Jiaotong University, Xi’an, Shaanxi, 710061, China. Fengjie Gao and Hairong He contributed equally to this study.

**Keywords:** network meta-analysis, panic disorder, CBT, combined therapy

## Abstract

**Objective:** To compare 22 oral drugs, cognitive behavior therapy (CBT), and their combination treatments for the acute treatment of adults with panic disorder in terms of remission rate and acceptability.

**Design:** Systematic review and network meta-analysis

**Data sources:** PubMed, Web of Science, Cochrane Central Register of Controlled Trials, and Embase databases from their inception up to May 26, 2019.

**Study selection:** Randomized controlled clinical trials (RCTs) of any oral drugs, CBT, CBT combined with any drug, or placebo in the acute treatment of adults with panic disorder diagnosed according to standard operationalized criteria. The primary outcomes were efficacy (remission rate) and acceptability (treatment discontinuations due to any cause).

**Results:** We identified 6585 reports that included 68 full-text RCTs involving 11101 patients. In terms of efficacy, 13 (68%) of 19 interventions were associated with higher remission rates than those of for placebo, with ORs ranging from 2.1 (95% credible interval [CrI] = 1.1 to 4.0) for sertraline to 13 (CrI = 4.5 to 44) for CBT combined with any drug. Regarding acceptability, alprazolam, imipramine, and etizolam were associated with lower dropout rates, with ORs ranging from 0.23 (CrI = 0.15 to 0.33) for alprazolam to 0.076 (CrI = 0.0021 to 0.77) for etizolam. Most of the differences between the other interventions were unclear. In head-to-head analyses, CBT combined with any drug was more effective than the other interventions, but it was no associated with an improvement in acceptability (OR = 0.12 to 0.219).

**Conclusions:** CBT combined with any drug was more effective than the other interventions analyzed in this study. CBT alone did not differ significantly from other drugs alone. We found that most drugs are effective against panic disorder, but they exhibit different acceptability and tolerability profiles.

**What is already known on this topic:** Some randomized controlled trials indicated that combined therapy are more effective than drug alone or cognitive behavior therapy alone. Nevertheless, drug monotherapy, cognitive behavior therapy, and combination therapy had never been compared in a comprehensive network meta-analysis. With this evidence gap remaining, the availability of reliable evidence of the relative merits of multiple interventions is essential to ensuring that clinicians choose the best option for each individual patient.

**What this study adds:** This network meta-analysis is the first to evaluate the relative merits of cognitive behavior therapy, cognitive behavior therapy combined with any drug, and 22 oral drugs in the field of panic disorder. The results showed a significant higher remission rates for cognitive behavior therapy combined with any drug than those of the other interventions. Cognitive behavior therapy alone was superior to placebo but did not differ significantly from other active drugs. Additionally, paroxetine, venlafaxine, fluoxetine, sertraline, and clonazepam are more recommendable than the other drugs based on their remission rates, scores on panic-symptoms scales, and acceptability.

## Introduction

Panic disorder (PD) is characterized by repeated, unexpected panic attacks in which at least 4 of 13 characteristic symptoms (e.g., racing heart rate, chest pain, and the fear of collapse) are experienced. PD has a prevalence of 1.6-2.2% of the world population.^1^ About 25% of patients with PD also have agoraphobia, which is associated with an increased severity and worse outcome of PD.^2^ PD with or without agoraphobia is disabling and is associated with substantial functional morbidity and reduced quality of life, which imposes a burden on both healthcare systems and society as a whole. ^3-5^

Three types of intervention are recommended for treating PD patients: pharmacotherapy, psychotherapy, and combination therapy.^6^ Recent guidelines^6-8^ consider antidepressants—mainly selective serotonin-reuptake inhibitors (SSRIs)—as the first-line treatment for PD due to their adverse-effects profile being more favorable than those of monoamine oxidase inhibitors (MAOIs) and tricyclic antidepressants (TCAs).

However, a meta-analysis indicated that combined therapy or psychotherapy alone may be chosen as the first-line treatment for PD with or without agoraphobia.^9^ Among many competing psychotherapies, a recent Cochrane review and network meta-analysis^10^ found CBT to be the most efficacious. Moreover, CBT is the most widely studied and validated psychotherapeutic treatment for PD, and more than 325 studies on CBT have been reported on in recent years.^11^

While there are some reviews associated with the treatment of PD, to our knowledge CBT, drug monotherapy, and combination treatments have never been compared in a comprehensive network meta-analysis. The availability of reliable evidence of the relative merits of multiple interventions is essential to ensuring that clinicians choose the best option for each individual patient, and new RCTs reported in recent years should be included in further analyses.

We therefore conducted a network meta-analysis to inform clinical practice by comparing different drugs alone, CBT, and CBT combined with any active drug for the acute treatment of adults with PD with or without agoraphobia.

## Methods

### Search strategy and selection criteria

We searched the PubMed, Web of Science, Cochrane Central Register of Controlled Trials (CENTRAL), and Embase databases from their inception up to May 26, 2019 without language restriction. We used both free-text and Medical Subject Headings (MeSH) word-search strategies with the following keywords: panic disorder, panic*, agoraphobia (the search terms are available in the appendix). The species was limited to human, but there was no restriction on the interventions. The reference lists of identified articles, systematic reviews, meta-analyses, and pooled analyses were screened for additional studies.

We included all prescribed drugs we screened, CBT, and CBT combined with any active drug. We defined CBT as therapy with or without physiological components and containing both cognitive and behavioral therapy elements. We only applied this definition of CBT because it is the most widely studied, and it has been validated that the co-administration of cognitive and behavioral therapeutic components is superior to administering behavioral components alone.^12^ We included both individual and group therapies. The therapies could be administered face-to-face, in self-help format (e.g., a book, computer, or the Internet), or remotely (e.g., via telephone or video conferencing).

We included randomized parallel controlled studies comparing an active intervention with placebo or another intervention for the acute treatment of adults with PD. We excluded crossover RCTs and quasi-randomized controlled trials in which treatment assignment was decided using methods such as alternate days of the week. We included participants aged ≥18 years of either sex with a primary diagnosis of PD with or without agoraphobia according to systematic diagnostic criteria (DSM-III, DSM-III-R, DSM-IV, DSM-IV-TR, ICD-10, and CCMD-3), and excluded patients with treatment resistance or concomitant medical illnesses. A concurrent other psychiatric disorder was not applied as an exclusion criterion.

The following information was entered into an predefined Excel worksheet: first author, publication year, diagnostic criteria, sample size, age distribution, sex ratio, main inclusion criteria, registration information, sponsorship information, primary outcomes, length of the RCT, analysis population, population setting, approach used to address missing data, conducted sites, active intervention, control intervention, sample sizes of each groups, remission rate, response rate, panic-symptoms scale, endpoint scores on the panic-symptom scale (mean±SD values), the safety population, the number of patients who withdrew from the RCT for any reason, and the number of patients who withdrew from the RCT due to adverse effects.

Both fixed-dose and flexible-dose studies were included in our analysis. If multiple doses were used in one trial, we applied the maximum dose recommended in the guideline. Moreover, when a positive drug was used as a control, we selected a comparable dose of the researched drug.

We recorded all outcomes that occurred during 1–6 months. We defined acute treatment as 8 weeks of treatment for both the remission rate and acceptability analysis. If different time points during 1–6 months were reported for the included studies, we preferred the time point closest to 8 weeks.

The screening procedure applied to the searched studies consisted of the initial screening of RCTs and relevant reviews based on titles and abstracts, and the second screening of the downloaded full texts, with final checking of the reviews for missing articles. Two investigators independently screened the studies, extracted the relevant information, and assessed the risk of bias. Any disagreement was resolved by consensus between two investigators, or with the suggestion from a third one when necessary.

We attempted to obtain missing data using one of the following methods: estimating them from published figures, calculating them from available data according to a validated imputation method,^13^ or contacting the study authors to supplement the incomplete data of the included trials if author contact information was available.

### Outcomes

Our primary outcomes were efficacy (remission rate measured by the total number of patients who reached a satisfactory end state as defined by global judgement by the original investigators; Examples are no full-symptom panic attacks on the Panic and Anticipatory Anxiety Scale and a Clinical Global Impression Severity Scale score of 1 or 2) and acceptability (treatment discontinuation quantified as the proportion of patients who withdrew for any reason). Secondary outcomes were endpoint scores on panic-symptoms scales, the response rate expressing the number of patients who had a substantial improvement from baseline, as defined by original investigators (examples are ‘very much or much improved’ according to the Clinical Global Impression Change Scale and more than 40% reduction in the score of the Panic Disorder Severity Scale), and tolerability quantified as the proportion of patients who dropped out early because of adverse events. When more than one panic-symptoms scale was reported on in one paper, we applied preferences in the following order:^10^

- PDSS > PAS (Panic and Agoraphobia Scale) > ASI-R > ASI > ACQ > BSQ > other scales specific for PD.
- CGI-S > CGI-I > GAS > GAF > other global scales.
- FQ-ag > FQ-global > MI-AAL (Mobile Inventory for Agoraphobia-Avoidance-Alone) > MI-AAC (MI-Avoidance-Accompanied) > other scales specific for agoraphobia only.
- Panic frequency > panic severity > other scales specific for panic attacks only.

If both self-rated and observer-rated assessments were available for the chosen scale, we gave preference to the latter.

We prioritized the intention-to-treat (ITT) population in the efficacy analysis, and the secondary choice was the safety population if the ITT population was unavailable. The ITT population was defined as receiving drug therapy and having at least one outcome assessment. The safety population was defined as the population treated with the drug. The safety population was the first choice in acceptability analysis, and the randomized population could be used if the safety population was not reported.

### Patient involvement

No patients were involved in setting the research question or the outcome measures, nor were they involved in developing plans for design or implementation of the study. No patients were asked to advise on interpretation or writing up of results. There are no plans to disseminate the results of the research to study participants or the relevant patient community. We did not evaluate whether the studies included in the review had any patient involvement.

### Data analysis

We performed a network meta-analysis using the gemtc package^14, 15^ of R software, which is based on Bayesian theory. We estimated the odds ratio (OR) for dichotomous outcomes and the standardized mean differences (SMD, Cohen’s d) for continuous outcomes in the network meta-analysis. Binomial and normal likelihoods were used for dichotomous and continuous outcomes, respectively.

The study effect sizes were combined using a random-effects model. Forest plots were drawn in this model. We used the surface under the cumulative ranking curve (SUCRA) to rank the treatments for each outcome. In addition, we quantified the heterogeneity using the I^2^ statistic, where an I^2^ value exceeding 50% indicates substantial heterogeneity.^16^

We evaluated consistency using node splitting, which separates evidence on a particular comparison (node) into direct and indirect, and can be applied to networks where trial-level data are available.^17^

We applied Stata software (version 15.0) to analyze loop inconsistency and publication bias, and to draw network diagrams. Loop inconsistency refers to an inconsistency test based on variety of measure that constitute a ‘ring’, and is generally considered to be the basis for inconsistency.^18^ Publication bias was evaluated using funnel plots. The horizontal axis of a funnel plot indicates the difference between the effect of one paired comparison in a study and the combined effect of many similar comparisons, while the vertical axis normally shows the standard error of the effect size. The presence of symmetry about the zero line of a funnel plot indicates the absence of publication bias. No data points will be close to the horizontal axis if no small-sample effects are present. It is necessary to calculate study effect estimators for different comparisons in order to evaluate whether there are small-sample effects in the intervention network.^19^

### Quality assessment

We utilized the Cochrane Handbook for Systematic Reviews of Interventions in Review Manager software (version 5.3) to assess the risk of bias in the included studies. This assessment comprised the following seven items: randomization method, concealment of allocation, blinding of outcome assessors, blinding of study personnel and participants, incomplete outcome data, selective outcome reporting, and other sources of bias. The study quality was divided into the following three categories: (1) low-risk studies had no high-risk items and fewer than three items of unclear risk, (2) medium-risk studies had one high-risk item or more than four items of unclear risk, and (3) high-risk studies covered all other situations.^20^

## Results

### Description of included studies

Database searching identified 6585 records, with 5849 remaining after excluding duplicates. A further 5642 reports were excluded based on their titles and abstracts, and so 207 full-text articles were retrieved. Screening these articles yielded 68 full-text articles involving 11101 patients that compared 24 interventions or placebo (Figure S1). The mean study sample size was 184. 8086 patients were randomly assigned to an active intervention and 4466 were randomly assigned to placebo. The mean age of the patients was 36.13 years, and females comprised 58.66% of them. Most (n = 47, 69.12%) of the 68 studies were placebo-controlled trials, and 21 (30.88%) assigned patients to 3 or 4 groups. The studies included 32 (47.06%) multicenter studies, 34 (50%) included outpatients only, and 41 (60.29%) received external funding. Twenty-eight (41.18%) of the 68 studies recruited patients from North America, 24 (35.29%) recruited them from Europe, 5 (7.35%) recruited them from Asia, and 11 (16.18%) recruited them from other regions. The response rate was 65.01%, and the rate of all-cause discontinuation was 24.85%.

After screening, we analyzed all prescribed drugs we found, which comprised the following 22 oral drugs:

- SSRIs: fluoxetine, fluvoxamine, sertraline, paroxetine, citalopram, and escitalopram.
- TCAs: clomipramine, imipramine, maprotiline, and desipramine.
- MAOIs: moclobemide and brofaromine.
- Serotonin-noradrenaline-reuptake inhibitors (SNRIs): venlafaxine.
- Benzodiazepines (BZDs): alprazolam, clonazepam, lorazepam, etizolam, and adizolam.
- Noradrenergic and specific serotonergic antidepressants: mirtazapine.
- Noradrenergic-reuptake inhibitors: reboxetine.
- Others: ritanserin and buspirone.

The drugs that were combined with CBT were sertraline, fluvoxamine, moclobemide, and imipramine. The basic characteristics of all of the studies are summarized in Table S1. The overall risk of bias was high in 19 (27.9%) studies, moderate in 38 (55.9%), and low in 11 (16.2%) (Figure S2).

### Effect of interventions

The networks of eligible comparisons of the remission rate and acceptability are shown in Figure 1A and B, respectively. The remission rates for desipramine and brofaromine were not compared with placebo, which the acceptability of maprotiline and lorazepam were not directly compared with placebo. More details of pairwise meta-analysis are provided in Table S2.

Table 1 shows the results of the network meta-analysis of primary outcomes. In terms of remission rate (involving 46 RCTs and 8551 patients), 13 (68%) of 19 interventions were associated with higher remission rates than those for placebo, with ORs ranging from 2.1 (95% credible interval [CrI] = 1.1 to 4.0) for sertraline to 13 (CrI = 4.5 to 44) for CBT combined with any drug. In contrast, the remission rates for adinazolam, citalopram, lorazepam, moclobemide, desipramine, and reboxetine were lower than those for placebo. In terms of acceptability (involving 61 RCTs and 10761 patients), alprazolam, imipramine, and etizolam were associated with lower dropout rates due to any reason, with ORs ranging from 0.23 (CrI = 0.15 to 0.33) for alprazolam to 0.076 (CrI = 0.0021 to 0.77) for etizolam among 21 interventions. Most of the differences between the remaining interventions were unclear.

**Table 1.**
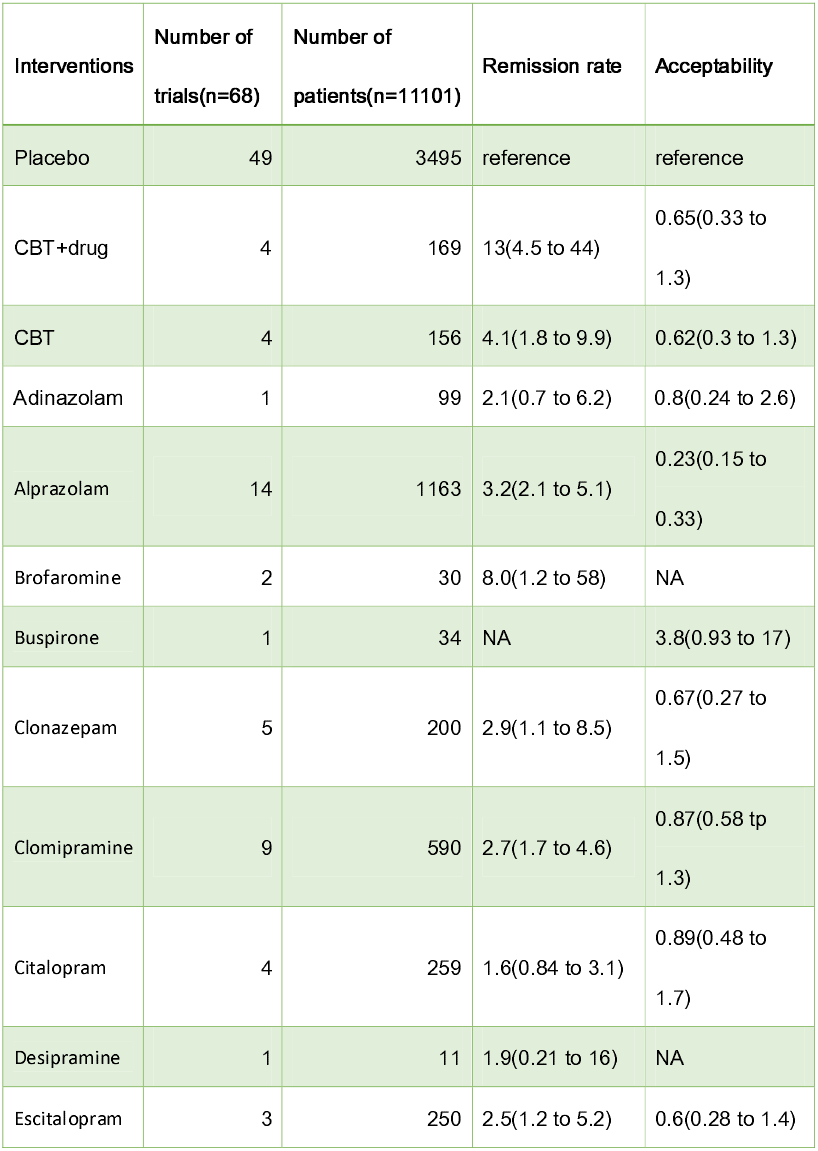
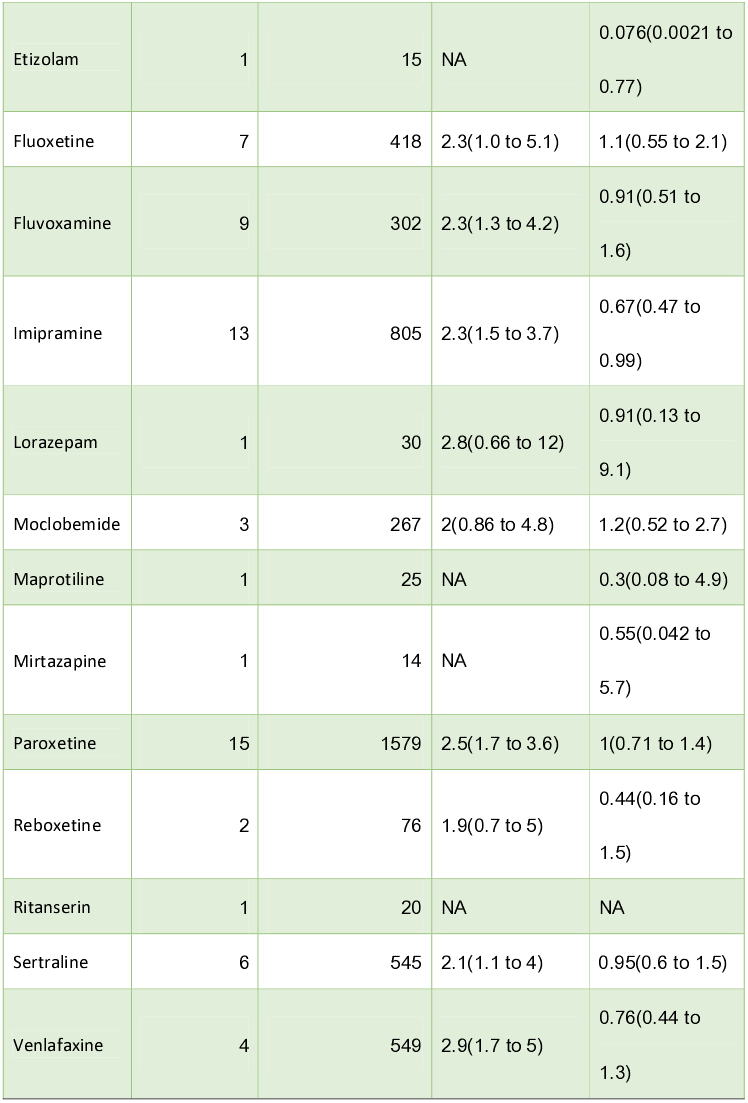
Treatment efficacy and acceptability compared with placebo. Data in parentheses are 95% credible intervals. CBT=cognitive behavior therapy. NA= not available.

Secondary outcomes were reported less frequently. Data regarding the original defined response rate were available for 34 studies involving 6193 patients. Ten (66%) of 15 interventions produced higher response rates than those for placebo, with ORs ranging from 2.1 (CrI = 1.1 to 4.0) for clomipramine to 10 (CrI = 4.4 to 25.0) for CBT combined with any drug (Figure 2A). However, the response rates for buspirone, citalopram, fluoxetine, moclobemide, and ritanserin did not differ significantly from those for placebo.

Results for panic symptoms (22 interventions) were reported for 35 studies (53%) involving 4420 patients. Paroxetine and clonazepam significantly reduced the panic symptoms compared with placebo, with SMDs of –1.7 (CrI **=** –2.8 to –0.67) and –2.2 (CrI **=** –3.8 to –0.62), respectively. The panic symptoms did not differ significantly between the other interventions and placebo (Figure 2B).

The dropout rates due to adverse events were reported for 38 studies involving 8967 patients. Clomipramine, fluvoxamine, and imipramine were associated with higher dropout rates than those for placebo, with ORs ranging from 2.2 (CrI = 1.1 to 4.4) for clomipramine to 2.9 (CrI = 1.2 to 6.9) for fluvoxamine. The dropout rates of the other interventions did not differ significantly from those for placebo (Figure 2C).

We also performed head-to-head analyses to assess the differences between interventions. Figure 3 shows the results for the primary outcomes. CBT combined with any drug was more effective than the other interventions, with ORs ranging from 0.12 to 0.219. In terms of acceptability, alprazolam and etizolam were associated with lower dropout rates than the other interventions, with ORs ranging from 1.9 to 17. In contrast, buspirone had higher dropout rates than the other interventions, with ORs ranging from 0.019 to 0.2. The differences between the remaining interventions were unclear.

### Inconsistency and publication bias

The results from the direct and indirect comparisons of remission rates were inconsistent for fluvoxamine versus CBT, imipramine versus clomipramine, paroxetine versus clomipramine, placebo versus imipramine, and placebo versus moclobemide. The results from direct and indirect comparisons of acceptability were inconsistent for fluvoxamine versus CBT, reboxetine versus paroxetine, and reboxetine versus placebo.

Our analysis of loop inconsistency revealed that 2 (8%) of 25 loops were inconsistent for analyzing the remission rate, and no loops were inconsistent for tolerability (Figure S3). No loops were found for acceptability and another two secondary outcomes in this analysis because insufficient observations were made in many of the included studies, such as for the CBT group. This means that there was no strong evidence of inconsistency for the explored outcomes, but the statistical power of this analysis is expected to be low.

The funnel plots were roughly symmetrical for each outcome, which indicated the absence of obvious publication bias. In addition, there was little evidence of small-sample effects for each outcome (Figure S4).

## Discussion

This network meta-analysis included 68 RCTs with 11101 patients comparing 1 of 24 interventions with another intervention or placebo. To our knowledge, this is the most comprehensive network meta-analysis of acute treatments for PD with or without agoraphobia, because it involved CBT combined with pharmacotherapy, CBT alone, and 22 oral drugs.

We found that most drugs alone, CBT, and CBT combined with any drug are more effective than placebo, with combination therapy being especially efficacious. To make our results more robust for informing clinical judgements, we focused on head-to-head analyses for the primary outcomes and found that CBT combined with any drug was more effective than the other interventions. The combined treatment of antidepressants and psychotherapy (behavior, CBT, or other types) was suggested to be a suitable first-line treatment for PD in a meta-analysis that included 21 studies with 1700 patients.^9^ In three RCTs, researchers combined CBT with sertraline, fluvoxamine, or imipramine, and found combination therapy to be superior to placebo or the drug alone.^21-23^ The present review was more comprehensive in including more RCTs, and it produced very similar results when the psychotherapy was restricted in CBT. The current findings indicate that combined therapy may be regarded as the first-choice intervention for the acute treatment of PD with or without agoraphobia.

CBT is effective for PD in many previous studies.^24-26^ Our results also shows that CBT was superior to placebo for remission rate. However, there is scarce evidence of the relative merits of CBT and pharmacotherapy. A meta-analysis published in 1995 indicated that CBT yield larger effect sizes than antidepressants or benzodiazepines.^27^ As new evidence, our results shows no significant differences between CBT and drugs for both remission rate and acceptability based on the head-to-head comparisons. Moreover, the fact that CBT is well tolerate due to absence of adverse events makes it recommendable for PD in clinical practice.

Guidelines from the UK National Institute of Clinical Excellence^6^ suggest that SSRIs should be used as first-line drugs. Our results strongly support this, with SSRIs including fluoxetine, fluvoxamine, sertraline, paroxetine and escitalopram being more effective than placebo. Among these drugs, paroxetine can significantly reduce scores on panic-symptoms scales compared with placebo, as evaluated by the SMD, whose use can greatly reduce heterogeneity.^28^ However, the present study produced the inconsistent result that citalopram had a smaller effect size than placebo. It should also be noted that fluvoxamine was tolerated less well than placebo.

Although treatment guidelines favor SSRIs over BZDs based on the belief that BZDs are associated with more risks of adverse effects than SSRIs,^29^ many authors pointed out that these risks have been overestimated and there are no high-quality studies demonstrating these risks.^30-33^ For this controversial question, our results shows that tolerability for BZDs including alprazolam and clonazepam were not significantly different with that for placebo. Moreover, in the head-to-head comparisons of all available interventions, we found that the dropout rate due to any reason for alprazolam was lower than those for many other drugs, and there are no significant differences between alprazolam, clonazepam and individual SSRIs in remission rate. These findings may support the guideline makers for PD to overcome the bias against BZDs and refrain from the unjustifiable avoidance of BZDs.

In addition, SNRIs such as venlafaxine, and TCAs including clomipramine and imipramine are more effective than placebo. It is noteworthy that clomipramine and imipramine were tolerated less well than placebo in this analysis, which may be due to severe adverse effects such as anticholinergicity.^34^ In contrast, the lower remission or response rates of adinazolam, lorazepam, moclobemide, reboxetine, desipramine, ritanserin, and buspirone make them less favorable. Furthermore, some drugs such as duloxetine and agomelatine are effective in patients with PD, as confirmed in some open-label trials,^35, 36^ but they were not included in the present network meta-analysis due to none of our screened RCTs including them. We hope that new evidence will be obtained to fill these gaps and thereby validate the present results.

CBT combined with pharmacotherapy may be used as the first-choice intervention for PD based on the present results, but its implementation would require policy changes to increase access to a sufficient number of trained therapists.^37^ Considering this disadvantage, pharmacotherapy might be preferable for many prescribers. According to the present primary and secondary outcomes, we suggest that paroxetine, venlafaxine, fluoxetine, sertraline, alprazolam, and clonazepam can be used as first-line drugs for PD. These results may inform clinicians, guideline makers, and patients about the relative merits of CBT combined with any drug, CBT, and many drugs used in PD with or without agoraphobia.

### Limitations of this study

Firstly, the small number of studies for many interventions, such as brofaromine, buspirone, mirtazapine, lorazepam, ritanserin, desipramine, and maprotiline, rendered the further explorations of sensitivity analysis and subgroup analysis. Secondly, the finding of larger effect sizes for smaller trials may be attributable to small-sample effects even though we found little evidence of small-sample effects in the funnel plots. For example, the analysis of etizolam was based on only 1 study with a sample size of 15, which may have exaggerate its effect. Thirdly, the placebo response of each intervention threatens the transitivity of assumptions based on a network meta-analysis.^38^ Although our findings are subjected by these limitations and insufficient high-quality RCTs included, combined therapy is also a potential and highly effective treatment towards PD, which is notably needed to be further researched in the future. We hope high-quality studies could be conducted to address these limitations and thereby improve the ability to interpret the present results. Clinicians should additionally remember that our results represent the average effects found in the included studies, and neglect the effects of age, sex, severity, and duration. This means that individual patients encountered in clinical practice may exhibit completely different responses.

Most of the RCTs included in this study were only of moderate or low quality, which may have been due to the unclear risks of randomization and allocation concealment. In addition, full patient blinding cannot be implemented for CBT and CBT-with-drug groups, but observer blinding can be achieved.

There are several potential explanations for the present heterogeneity. Firstly, the different type of interventions applied to PD may inevitably cause heterogeneity. Secondly, we included patients complicated with other psychiatric disorders, such as major depression, social phobia, specific phobia, and generalized anxiety disorder. The comorbidity of different psychiatric disease may induce heterogeneity, but we could not exclude these studies because such comorbidity is very common in patients with PD.^39^ Thirdly, the duration of treatment and drug doses differed between the included studies; nevertheless, we restricted these variations in the inclusion criteria. All of these factors should be considered carefully when interpreting the results of this study.

## Conclusion

The present analysis of 24 interventions found that the combination of CBT and any drug may possibly be used as the first-line intervention for PD based on its significant remission rate. Moreover, our data indicate that there is no significant differences between CBT and drugs for both remission rate and acceptability. Among the analyzed drugs, our results shows that many drugs are effective, for instance paroxetine, venlafaxine, fluoxetine, sertraline, alprazolam, and clonazepam, but they do exhibit variable acceptability and tolerability profiles. These findings may assist health-care providers to choose the most suitable treatment for individual patients with PD.

## Data Availability

All data are available from corespondence author.

## Contributors

Wei Wang and Xiancang Ma contributed equally to the work. Wei Wang and Xiancang Ma contributed to the study concept and study design. Fengjie Gao and Hairong He performed statistical analysis and data interpretation. Fengjie Gao, Hairong He, and Feng Zhu performed literature research and data extraction. Jian Yang and Yajuan Fan were responsible for the quality control of data and algorithms. Fengjie Gao, Hairong He, and Xiancang Ma contributed to the writing of the manuscript. All authors contributed to writing of the manuscript and approved the final version.

## Ethical approval

Not required.

## Data sharing

No additional data available.

## Transparency

The lead authors affirm that the manuscript is an honest, accurate, and transparent account of the study being reported; that no important aspects of the study as planned have been explained.

This is an Open Access article distributed in accordance with the Creative Commons Attribution Non Commercial (CC BY-NC 4.0) license, which permits others to distribute, remix, adapt, build upon this work non-commercially, and license their derivative works on different terms, provided the original work is properly cited and the use is non-commercial. See: http://creativecommons.org/licenses/by-nc/4.0/.

---

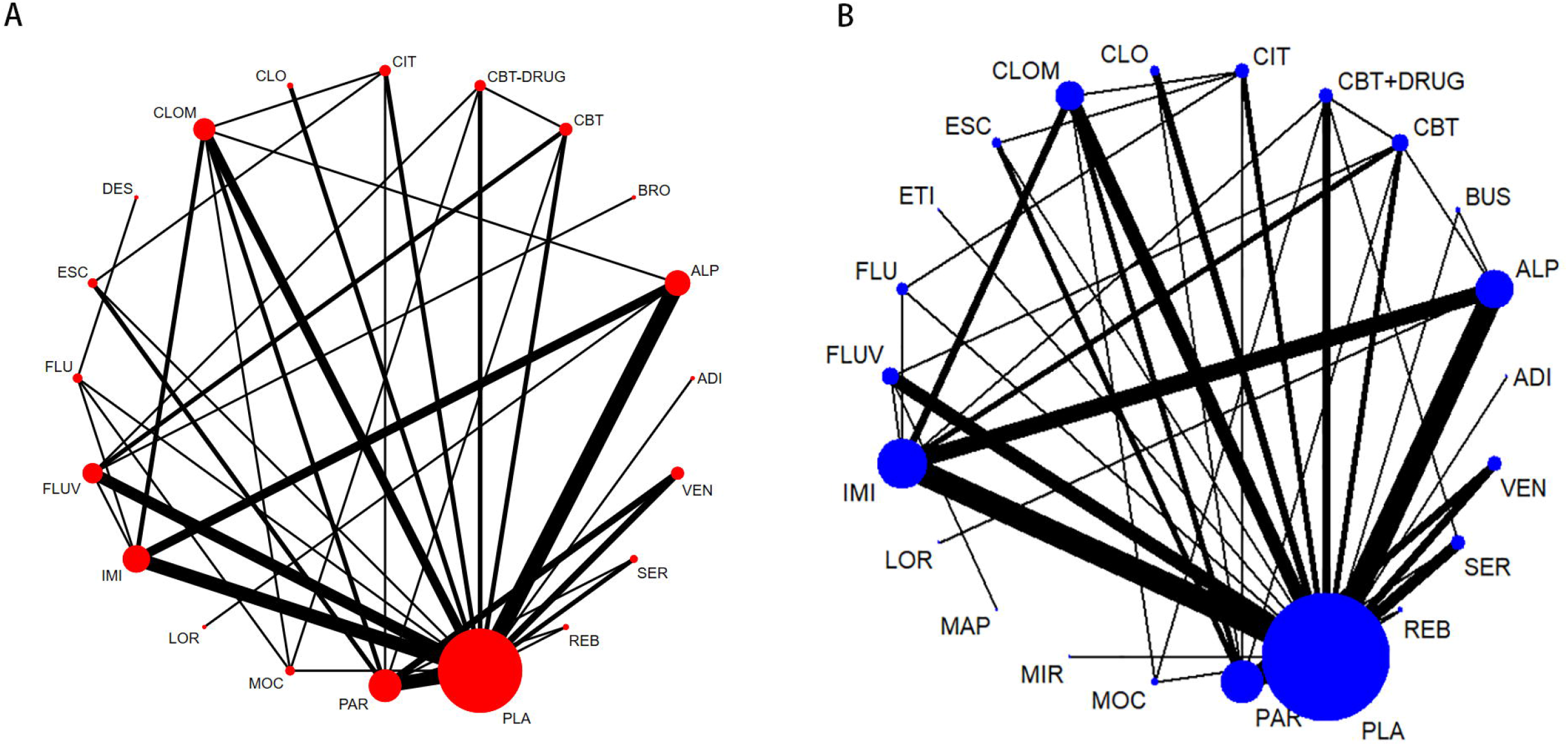

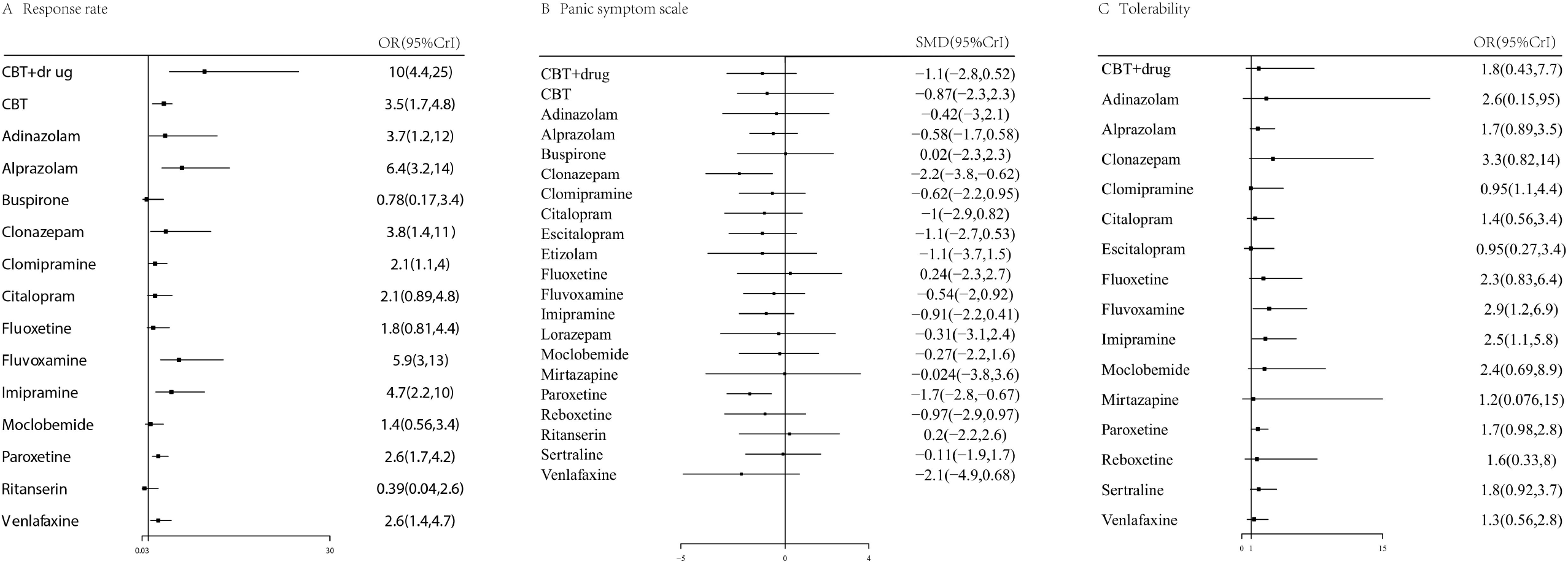

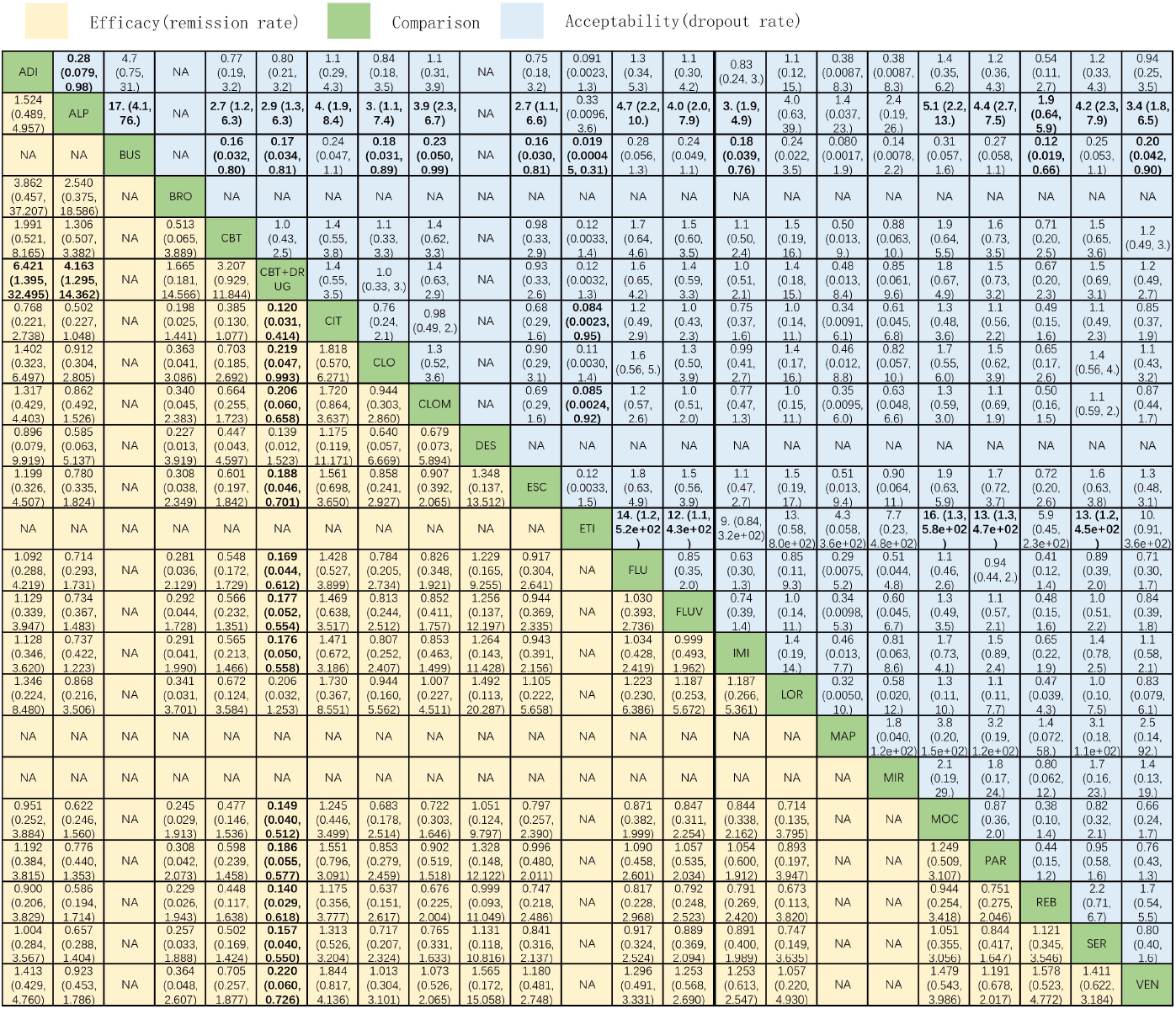

